# Uncovering Survivorship Bias in Longitudinal Mental Health Surveys During the COVID-19 Pandemic

**DOI:** 10.1101/2021.01.28.21250694

**Authors:** M. Czeisler, J. Wiley, C. Czeisler, S. Rajaratnam, M. Howard

## Abstract

**Aims:** Markedly elevated adverse mental health symptoms were widely observed early in the coronavirus disease 2019 (COVID-19) pandemic. Unlike the U.S., where cross-sectional data indicate anxiety and depression symptoms have remained elevated, such symptoms reportedly declined in the U.K., according to analysis of repeated measures from a largescale longitudinal study. However, nearly 40% of U.K. respondents (those who did not complete multiple follow-up surveys) were excluded from analysis, suggesting that survivorship bias might partially explain this discrepancy. We therefore sought to assess survivorship bias among participants in our longitudinal survey study as part of The COVID-19 Outbreak Public Evaluation (COPE) Initiative.

**Methods:** Survivorship bias was assessed 4,039 U.S. respondents who completed surveys including the assessment of mental health as part of The COPE Initiative in April 2020 and were invited to complete follow-up surveys. Participants completed validated screening instruments for symptoms of anxiety, depression, and insomnia. Survivorship bias was assessed for (1) demographic differences in follow-up survey participation, (2) differences in initial adverse mental health symptom prevalences adjusted for demographic factors, and (3) differences in follow-up survey participation based on mental health experiences adjusted for demographic factors.

**Results:** Adjusting for demographics, individuals who completed only one or two out of four surveys had higher prevalences of anxiety and depression symptoms in April 2020 (e.g., one-survey *versus* four-survey, anxiety symptoms, adjusted prevalence ratio [aPR]: 1.30, 95% confidence interval [CI]: 1.08-1.55, *P*=0.0045; depression symptoms, aPR: 1.43, 95% CI: 1.17-1.75, *P*=0.00052). Moreover, individuals who experienced incident anxiety or depression symptoms had higher odds of not completing follow-up surveys (adjusted odds ratio [aOR]: 1.68, 95% CI: 1.22-2.31, *P*=0.0015, aOR: 1.56, 95% CI: 1.15-2.12, *P*=0.0046, respectively).

**Conclusions:** Our findings revealed significant survivorship bias among longitudinal survey respondents, indicating that restricting analytic samples to only respondents who provide repeated assessments in longitudinal survey studies could lead to overly optimistic interpretations of mental health trends over time. Cross-sectional or planned missing data designs may provide more accurate estimates of population-level adverse mental health symptom prevalences than longitudinal surveys.

## Introduction

Studies have documented acutely elevated prevalences of adverse mental health symptoms during the early months of the coronavirus disease 2019 (COVID-19) pandemic compared with pre-pandemic data.^1-9^ Prevalences of clinically significant mental distress rose by approximately 40% in the U.K.,^1^ and prevalences of anxiety and depression symptoms more than tripled in the U.S.^2-4^ Analysis of longitudinal U.K. survey data recently suggested that those increased prevalences may have been transient, with anxiety and depression symptoms declining among participants who completed two-or-more follow-up measures between April and August 2020.^10^ However, those longitudinal data from repeat-responders are not consistent with cross-sectional U.S. survey data, which indicate that levels of adverse mental health symptoms have remained persistently elevated.^11,12^ As 38.5% of U.K. respondents were excluded from analysis because did not complete multiple follow-up surveys, we analysed data from U.S. adults invited to complete surveys over a comparable time interval to determine if survivorship bias could account for the discrepancy between the published data from U.S. and U.K. adults. This investigation has practical and theoretical implications. Reliable assessment of the prevalences of adverse mental health symptoms could affect planning and resource allocation for mental health support services during the COVID-19 pandemic.^13^ More broadly, given that survivorship bias has not previously been reported to affect largescale internet-based mental health surveys, this investigation may influence mental health study design and interpretation of ongoing studies and previously published papers.

While bias induced by demographic differences in follow-up survey participation may be reduced by poststratification weighting using population estimates,^14^ this strategy cannot account for survivorship bias. Survivorship bias can be problematic if individuals who make it past a selection process are different than those who do not. In the context of longitudinal mental health surveys, this could result from restricting an analytic sample to respondents who consistently participated in surveys, ignoring individuals who dropped out. If the people who dropped out (i.e., study non-survivors) were to have meaningfully different mental health than those who remain active study participants (i.e., study survivors), the resulting analytic sample would be non-representative.

Longitudinal studies have provided evidence of survivorship bias related to mental health within specific populations.^15-23^ For example, diagnosed depression has been associated with lower participation in follow-up surveys in parents and children^15,16^ and a naturalistic cohort on depression and anxiety,^17^ while assessment of three-year follow-up surveys in the Netherlands general population reported no association between mental health status at baseline and attrition.^18^ However, considerable effort was exerted to optimise participation, including a two-year initial contact and follow-up intervals, multiple attempts to recontact participants, and frequent contact between interviews. Other studies have that cancer survivors who completed surveys at multiple time points had higher health-related quality of life scores than those who completed surveys at a single timepoint^19^ and pregnant persons with psychological distress had higher odds of not completing follow-up surveys compared with pregnant persons without such distress.^20^ Additionally, non-participation in follow-up surveys has been associated with smoking and alcohol use among trauma patients,^21^ and with lower perceived oral healthcare-specific self-efficacy among patients with chronic periodontitis.^22^ Finally, of 294 women who presented at an emergency department following sexual assault, 136 (46%) could not be reached within 48 hours, and 233 (79%) did not participate in six-month follow-up.^23^ While anxiety and depression symptom ratings were attenuated in the analytic sample of 61 women who completed six-month follow-up surveys, women with higher rape-trauma-symptom scores were more likely to decline follow-up surveys. If survivorship bias existed in that study, generalizing data supporting declining adverse mental health levels from only those with lower initial rape-trauma-symptom scores could lead to an overly optimistic interpretation of mental health following sexual assault.

To our knowledge, survivorship bias assessment has not been described in general population longitudinal mental health internet-based survey data and is seldom addressed. As numerous studies have responded to the call for mental health research by launching longitudinal mental health survey studies, we undertook a robust assessment of potential survivorship bias in our longitudinal mental health survey study.

## Methods

### Study design

We conducted a retrospective analysis of U.S. participants in The COVID-19 Outbreak Public Evaluation (COPE) Initiative (www.thecopeinitiative.org).^24^ Internet-based surveys were administered through Qualtrics, LLC^24^ to 4,042 U.S. adults aged ≥18 years during April 2-8, 2020 (April-2020). The sample included 3,010 (74.5%) from across the U.S., plus additional respondents from New York City (n: 507 [12.5%]) and Los Angeles (n: 525 [13.0%]) to recruit participants from cities with different prevalences of SARS-CoV-2 during the early months of the pandemic.^26^ All respondents were invited to complete follow-up surveys during May 5-12, 2020 (May-2020) and June 24-30, 2020 (June-2020).^2^ Respondents who completed at least one of these follow-up surveys were also invited to complete surveys during August 28-September 6, 2020 (September-2020). Quota sampling and survey weighting (iterative proportional fitting with weights trimmed between 0.3 and 3.0, inclusive) were employed to improve sample representativeness by gender, age, and race/ethnicity using Census population estimates. Given that available Census data did not report on gender, for this analysis, Census data on sex was used to for weighting of dichotomized gender. One respondent who was inadvertently invited to and completed a September-2020 survey after not having participated in May-2020 or June-2020 surveys and two respondents who identified as “Other” gender were not included in this analysis.

Surveys contained demographic questions and assessed public attitudes and behaviours related to the pandemic and its mitigation, along with mental health symptoms. Validated screening instruments and modified questions from instruments were used. Among the mental health screening instruments were the 4-item Patient Health Questionnaire (PHQ),^27^ with subscales for assessment of anxiety (2-item Generalized Anxiety Disorder [GAD-2]) and depression (2-item PHQ [PHQ-2]) symptoms, and the 2-item Sleep Condition Indicator (SCI-02) for assessment of insomnia symptoms.^28^

### Statistical analysis

We explored whether potential mental health survivorship bias could be explained by: (1) demographic differences in repeated measures respondents (i.e., cross-sectional versus longitudinal respondents differ in their demographics, but mental health is similar among members of a demographic subgroup); or (2) differences within demographic subgroups. Demographic survey weighting could considerably reduce bias in the first, but not second scenario.

Potential demographic differences in survey retention were assessed using chi-square tests for differences between the percentages of respondents who completed one, two, there, or four surveys by gender, age group in years, combined race/ethnicity, education attainment, and 2019 household income. Potential differences in initial mental health were assessed using weighted Poisson regression models with robust standard error estimators to estimate prevalence ratios (PRs) and 95% confidence intervals (CIs) for April-2020 anxiety symptoms (≥3 out of 6 on the GAD-2 subscale of the PHQ-4), depression symptoms (≥3 out of 6 on the PHQ-2 subscale of the PHQ-4), and insomnia symptoms (≤2 out of 8 on the SCI-02) based on the number of completed surveys, both unadjusted and adjusted for gender, age group, race/ethnicity, education attainment, and 2019 household income. Next, to assess for potential differences in population estimates for prevalences of anxiety, depression, and insomnia symptoms in April 2020 using samples with differing retention over time, the April-2020 sample was separated into four groups: respondents who completed one, two, three, or four surveys through September 2020. Each group was separately weighted to improve representativeness of the U.S. population by gender, age, and race/ethnicity. Prevalences of anxiety, depression, and insomnia symptoms were estimated based on these demographically representative groups. Chi-square tests were used to assess for different point-estimates for prevalence of April-2020 anxiety, depression, and insomnia symptoms between groups.

To evaluate potential differences in trajectories of adverse mental health symptoms over time by number of completed surveys, prevalences of symptoms of anxiety, depression, and insomnia over two timepoints (April-2020 to May-2020 and April-2020 to June-2020) among respondents who completed all four surveys were compared with those who completed two total surveys (only April-2020 and May-2020 or only April-2020 and June-2020). Respondents who participated in all four surveys completed three of three follow-up surveys (100% retention rate), whereas respondents who participated in two surveys only completed one of three follow-up surveys (33% retention rate). Chi-square tests were used to assess for differences in initial (April-2020) prevalence between samples, and for differences over time (April-2020 *versus* May-2020) within each sample. Prevalence ratios were used to estimate differences in prevalences between subsamples over time.

Finally, to assess whether changes in mental health symptoms were associated with differential participation in follow-up surveys, weighted ordinal logistic regressions were used to estimate odds ratios for lower participation in in June-2020 and September-2020 surveys among respondents who completed April-2020 and May-2020 surveys based on symptoms of anxiety, depression, or insomnia reported in these two initial surveys. For each of these adverse mental health conditions over April-2020 and May-2020, respondents were categorized as having no symptoms at either timepoint, symptoms at both timepoints, incident symptoms in May-2020 after not having experienced symptoms in April-2020, or remitted symptoms in May-2020 after having experienced symptoms in April-2020. Odds ratios for lower participation in follow-up surveys were estimated with the dependent variables ordered as 0 (completed both follow-up surveys), 1 (completed one follow-up survey [either June-2020 or September-2020]), and 2 (completed neither follow-up survey). Odds were estimated both unadjusted and adjusted for gender, age group, race/ethnicity, education attainment, and 2019 household income.

### Study approval and informed consent

The Monash University Human Research Ethics Committee approved the study protocol. Participants provided electronic informed consent. Rounded weighted values are reported unless otherwise specified. Analyses were conducted in R (version 4.0.2; The R Foundation)^29^ with the R survey package (version 3.29)^30-32^ and Python (version 3.7.8).^33^

## Results

Overall, 4,042 of 6,548 (61.7%) eligible invited adults completed surveys during the first wave of The COVID-19 Outbreak Public Evaluation (COPE) Initiative, administered during April 2-8, 2020. Of 4,039 (99.9%) who were included in this analysis, 2,098 (52.0%) completed May-2020 surveys, 1,619 (40.0%) completed June-2020 surveys, and 1,151 (28.5%) completed September-2020 surveys. In total, 1,712 (42.4%) completed one survey, 725 (17.9%) completed two surveys, 663 (16.4%) completed three surveys, and 939 (23.2%) completed all four surveys (Table). By age, 76.0% of respondents aged 18-24 years completed one survey, whereas 7.3% completed three or four surveys. In contrast, just 12.1% of respondents aged ≥65 years completed one survey, compared with 72.5% who completed three of four surveys (*P*<2.20×10^−16^). By race/ethnicity, non-Hispanic White and non-Hispanic Asian respondents had the lowest prevalences of one-survey respondents (33.4% and 32.3%, respectively) and highest prevalences of four-survey respondents (29.7% and 23.9%), whereas non-Hispanic Black and Latinx respondents had the highest prevalences of one-survey respondents (65.7% and 60.7%, respectively) and lowest prevalences of four-survey respondents (8.6% and 10.1%); *P*<2.20×10^−16^. Percentage of completed surveys also increased with higher education attainment (e.g., one-survey, high school diploma or less: 52.9%, after bachelor’s degree: 33.1%, *P*<2.20×10^−16^) and higher 2019 household income (e.g., one-survey, USD <25,000: 51.0%, ≥100,000: 37.0%, *P*=1.68×^10-9^).

**Table.** Respondent characteristics, overall and by number of completed surveys

|  | Number of respondents |  |  |  | Number of completed surveys |  |  |  |  |  |  |  | Chi-Sq<br>*P* |
| --- | --- | --- | --- | --- | --- | --- | --- | --- | --- | --- | --- | --- | --- |
|  | Unweighted |  | Weighted |  | One |  | Two |  | Three |  | Four |  |  |
|  | n | (%) | n | (%) | n | (%) | n | (%) | n | (%) | n | (%) |  |
| **Total** | **4039** | **(100)** | **4039** | **(100)** | **1712** | **(42.4)** | **725** | **(17.9)** | **663** | **(16.4)** | **939** | **(23.2)** | - |
| **Gender** |  |  |  |  |  |  |  |  |  |  |  |  |  |
| Male | 1814 | (44.9) | 1986 | (49.2) | 872 | (43.9) | 329 | (16.6) | 307 | (15.5) | 477 | (24.0) | 0.032 |
| Female | 2225 | (55.1) | 2053 | (50.8) | 840 | (40.9) | 395 | (19.2) | 356 | (17.3) | 462 | (22.5) |  |
| **Age group in years** |  |  |  |  |  |  |  |  |  |  |  |  |  |
| 18-24 | 456 | (11.3) | 528 | (13.1) | 401 | (76.0) | 88 | (16.7) | 22 | (4.2) | 17 | (3.1) | <2.20×10<sup>-16*</sup> |
| 25-44 | 1335 | (33.1) | 1414 | (35.0) | 809 | (57.2) | 269 | (19.0) | 182 | (12.8) | 155 | (11.0) |  |
| 45-64 | 1420 | (35.2) | 1403 | (34.7) | 418 | (29.8) | 261 | (18.6) | 291 | (20.8) | 433 | (30.8) |  |
| ≥65 | 828 | (20.5) | 693 | (17.2) | 84 | (12.1) | 107 | (15.4) | 168 | (24.2) | 335 | (48.3) |  |
| **Race/ethnicity** |  |  |  |  |  |  |  |  |  |  |  |  |  |
| White, non-Hispanic | 2937 | (72.7) | 2575 | (63.7) | 860 | (33.4) | 459 | (17.8) | 491 | (19.1) | 765 | (29.7) | <2.20×10<sup>-16*</sup> |
| Black, non-Hispanic | 329 | (8.1) | 493 | (12.2) | 324 | (65.7) | 72 | (14.6) | 55 | (11.1) | 43 | (8.6) |  |
| Asian, non-Hispanic | 224 | (5.5) | 189 | (4.7) | 61 | (32.3) | 45 | (24.0) | 37 | (19.8) | 45 | (23.9) |  |
| Other, non-Hispanic | 126 | (3.1) | 122 | (3.0) | 66 | (54.4) | 22 | (18.5) | 13 | (11.0) | 20 | (16.1) |  |
| Latinx | 423 | (10.5) | 660 | (16.3) | 401 | (60.7) | 126 | (19.0) | 67 | (10.1) | 67 | (10.1) |  |
| **Education attainment** |  |  |  |  |  |  |  |  |  |  |  |  |  |
| ≤ High school diploma | 735 | (18.2) | 777 | (19.2) | 411 | (52.9) | 137 | (17.7) | 86 | (11.0) | 143 | (18.4) | <2.20×10<sup>-16*</sup> |
| College or some college | 2484 | (61.5) | 2473 | (61.2) | 1023 | (41.4) | 445 | (18.0) | 426 | (17.2) | 579 | (23.4) |  |
| > Bachelor's degree | 792 | (19.6) | 756 | (18.7) | 250 | (33.1) | 142 | (18.8) | 150 | (19.8) | 215 | (28.4) |  |
| Unknown | 28 | (0.7) | 34 | (0.8) | 28 | (83.9) | 1 | (2.7) | 2 | (5.4) | 3 | (8.0) |  |
| **2019 Household income (USD)** |  |  |  |  |  |  |  |  |  |  |  |  |  |
| <25,000 | 578 | (14.3) | 607 | (15.0) | 309 | (51.0) | 111 | (18.3) | 86 | (14.1) | 101 | (16.7) | 1.68×10<sup>-9*</sup> |
| 25,000-49,999 | 816 | (20.2) | 834 | (20.6) | 395 | (47.4) | 155 | (18.5) | 112 | (13.5) | 172 | (20.6) |  |
| 50,000-99,999 | 1291 | (32.0) | 1271 | (31.5) | 489 | (38.5) | 227 | (17.9) | 235 | (18.5) | 320 | (25.1) |  |
| ≥100,000 | 1156 | (28.6) | 1125 | (27.9) | 416 | (37.0) | 202 | (17.9) | 203 | (18.0) | 305 | (27.1) |  |
| Unknown | 198 | (4.9) | 202 | (5.0) | 103 | (51.0) | 31 | (15.1) | 27 | (13.3) | 42 | (20.6) |  |

USD = United States Dollars
\* $P < 0.025$ .
The "Other, non-Hispanic," category includes respondents who identified as not Hispanic or Latino and as more than one race or as American Indian or Alaska Native, Native Hawaiian or Pacific Islander, or Other.

Compared with respondents who completed all four surveys, those who completed only one or two surveys had higher prevalences of anxiety and depression symptoms in April-2020 surveys (Figure 1). Differences remained after adjusting for gender, age, race/ethnicity, education attainment, and 2019 household income among respondents (e.g., one-survey *versus* four-survey, anxiety symptoms, aPR: 1.30, 95% CI: 1.08-1.55, *P*=0.0045; depression symptoms, 1.43, 1.17-1.75, *P*=0.00052). Adjusted prevalence of insomnia symptoms in April-2020 was higher among individuals who completed only one survey compared with those who completed all four surveys (aPR: 1.33, 95% CI: 1.09-1.62, *P*=0.0045). Prevalence estimates for April-2020 adverse mental health symptoms among groups of respondents who completed one, two, three, or four surveys—each separately weighted to improve group representativeness of the U.S. population by gender, age, and race/ethnicity—revealed that estimates for anxiety and depression symptoms based on respondents who completed only one or two surveys were higher than those for respondents who completed three or four surveys (e.g., one-survey *versus* four-survey, anxiety symptoms: 25.5% *versus* 19.8%, *P*=2.50×10^−5^; depression symptoms: 24.1% *versus* 15.6%, *P*=1.91×10^−10^) (Figure 2). Prevalence estimates for these symptoms were similar between one- and two-survey respondents, and between three- and four-survey respondents. Estimates for insomnia symptoms were greater among respondents who completed one survey compared with those who completed multiple surveys (e.g., one-survey *versus* two-survey: 19.9% *versus* 16.1%, *P*=0.0097), but not among any respondents’ groups who completed multiple surveys.

**Figure 1.**
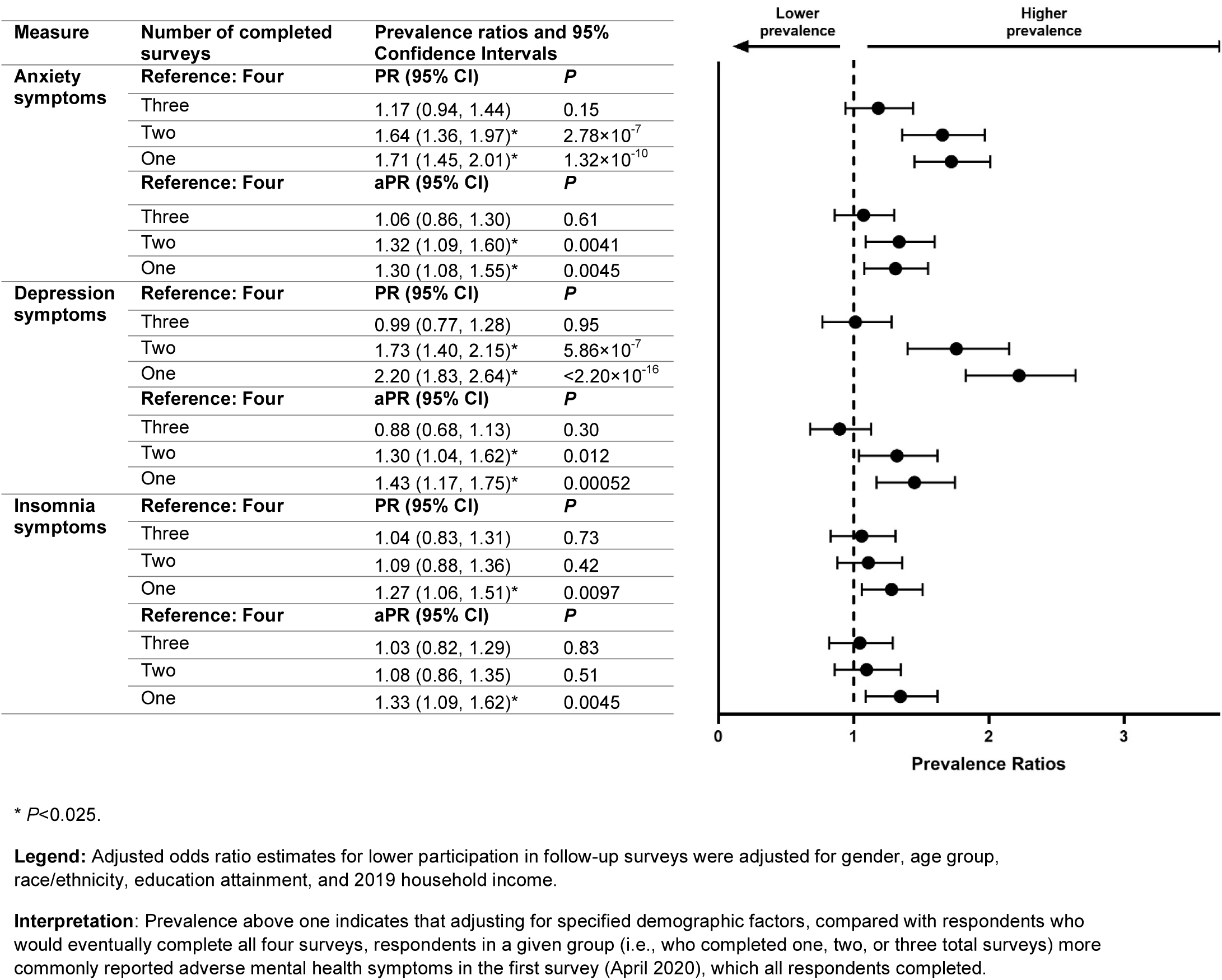
Adjusted prevalence ratios of anxiety, depression, and insomnia symptoms in April 2020 by number of completed surveys.

**Figure 2.**
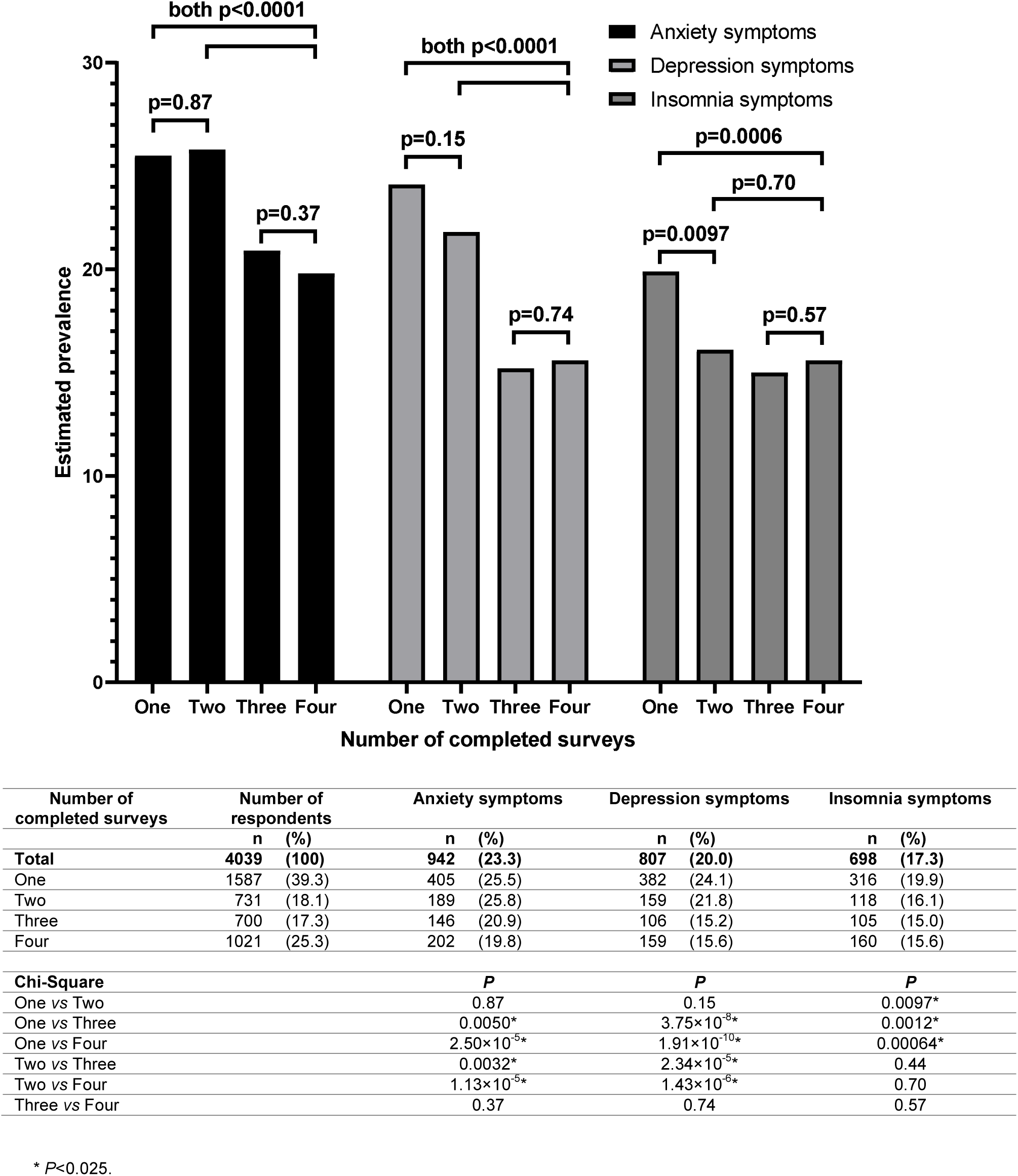
Estimated prevalences of symptoms of anxiety, depression, and insomnia in April 2020 based on total number of completes surveys, with each group weighted to population estimates for gender, age, and race/ethnicity.

In the comparison of adverse mental health symptom prevalences among respondents who completed only two surveys versus those who completed all four surveys (n: 939), both two-survey groups (April-2020 and May-2020 only [April-and-May; n: 584], April-2020 and June-2020 only [April-and-June; n: 141]) started with higher April-2020 prevalences of anxiety and depression symptoms (April-and-May, anxiety symptoms PR: 1.57, depression symptoms PR: 1.66; April-and-June: 1.91 and 2.02, respectively), and the prevalence ratios increased for the second completed surveys (April-and-May: 2.15 and 1.99, respectively; April-and-June: 2.55 and 2.33, respectively) (Figure 3). The prevalence of anxiety symptoms among April-and-May and April-and-June two-survey respondents was similar between surveys (April-and-May: 25.8% and 28.6%, respectively, *P*=0.19; April-and-June: 31.3% and 33.9%, respectively, *P*=0.57), whereas the prevalence of anxiety symptoms in four-survey respondents decreased over these intervals (April-and-May: 16.4% and 13.3%, *P*=0.012; April-and-June: 16.4% and 11.1%, *P*=1.11×10^−5^). The prevalence of depression symptoms increased among April-and-May two-survey respondents (21.4% and 27.5%, respectively, *P*=0.0017), but not among four-survey respondents (12.9% and 13.8%, *P*=0.45).

**Figure 3.**
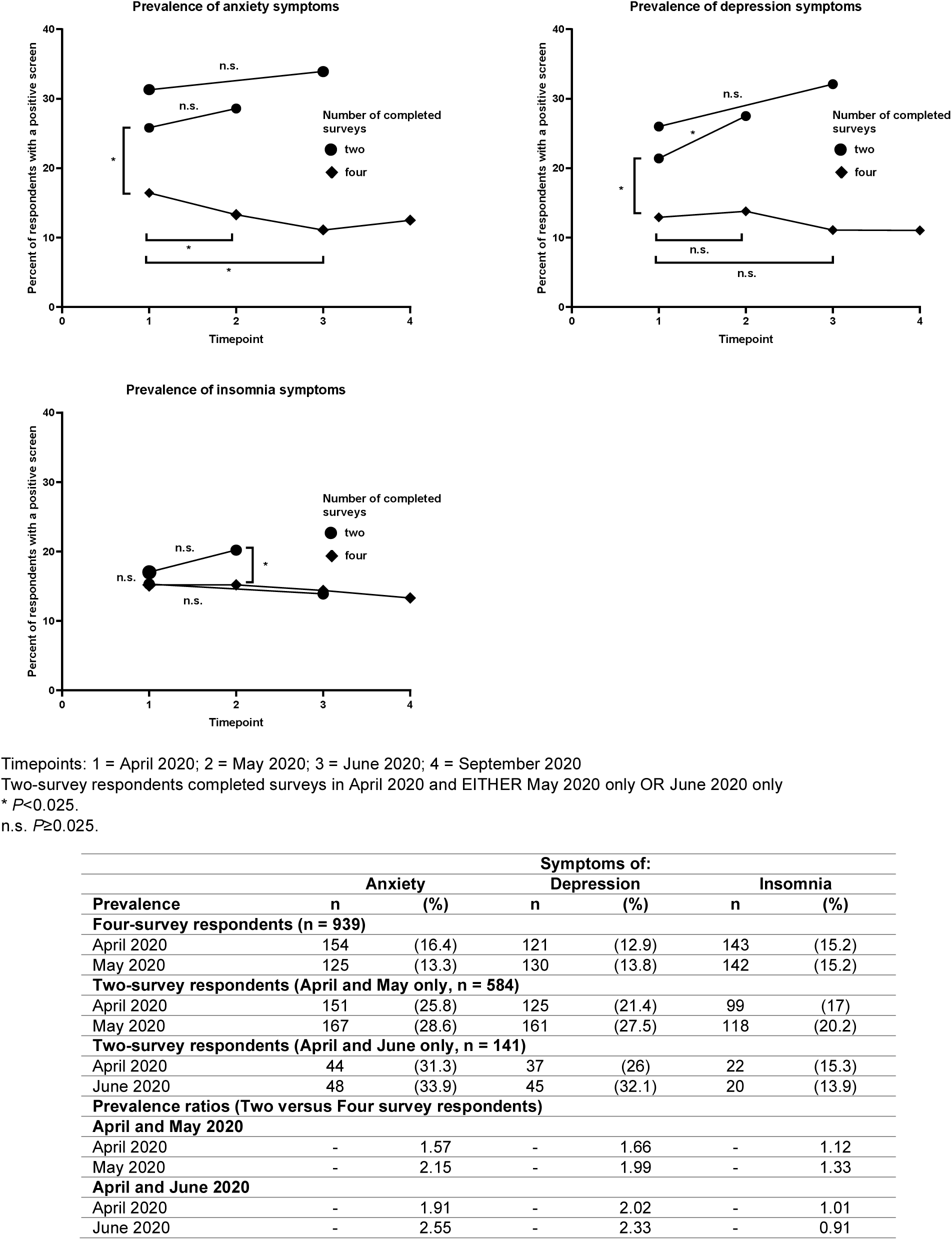
Longitudinal comparisons of anxiety and depression symptom prevalences by number of repeated measures.

Analysis of respondents who completed April-2020 and May-2020 surveys revealed that, compared with individuals who did not experience anxiety or depression symptoms during these initial surveys, those who experienced incident anxiety or depression symptoms had increased odds of lower participation in future follow-up surveys (i.e., June-2020 and September-2020) (Figure 4). Individuals who experienced anxiety symptoms and depression symptoms in May-2020 after not having done so in April-2020 had 1.68-times (1.22-2.31, *P*=0.0015) and 1.56-times (1.15-2.12, *P*=0.0046) increased odds, respectively, of lower participation in June-2020 and September-2020 surveys. Odds of follow-up survey participation was similar by insomnia symptoms, or among those who experienced (1) remission of anxiety or depression symptoms or (2) persistent anxiety or depression symptoms compared with those who did not experience these symptoms in April-2020 or May-2020.

**Figure 4.**
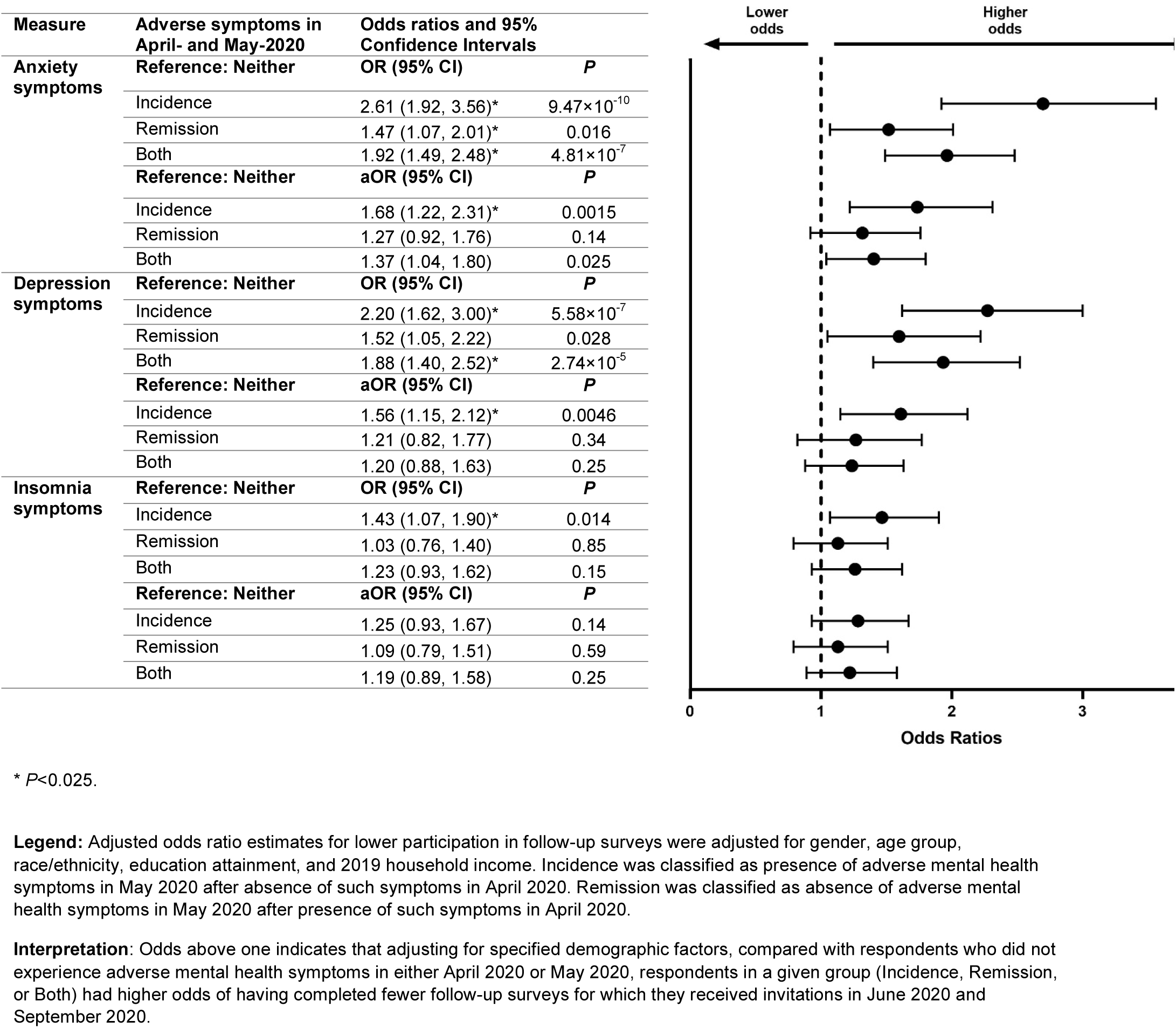
Odds of lower participation in follow-up surveys based on mental health in earlier surveys.

## Discussion

Analysis of mental health among survey respondents based on their participation in follow-up surveys revealed considerable survivorship bias related to: (1) demographic differences in survey retention; (2) differences in initial mental health, adjusted for gender, age, race/ethnicity, education, and income; and (3) higher odds of lower participation in follow-up surveys among respondents who experienced worsened mental health over time. The first of these forms of survivorship bias can be reduced by the application of poststratification weights. The second of these forms of survivorship bias precludes use of a longitudinal sample alone to estimate population prevalences of adverse mental health symptoms. However, simultaneous collection of cross-sectional data from representative samples of independent participants could inform strategies to mitigate differences in initial prevalences of adverse mental health symptoms, which could include adjustment for baseline differences in mental health between cross-sectional versus longitudinal respondents. The third of these forms of survivorship bias is most challenging to account for given the unknown trajectories of respondents who do not consistently participate in follow-up surveys. Recognition that individuals who experienced incident anxiety or depression symptoms had higher odds of not completing follow-up surveys reveals the hazard of overlooking this form of survivorship bias and should temper conclusions about trends of anxiety and depression symptoms in longitudinal mental health survey respondents, especially as generalizing from repeated survey administration among longitudinal respondents without addressing these biases could lead to potentially erroneous conclusions (e.g., that adverse mental health symptom prevalences in a population are improving over time).

Understanding strengths and limitations of study approaches should inform the design and interpretation of findings.^34^ Longitudinal studies have advantages, including increased power to detect causal pathways and mediating factors, reduced reliance on recall bias, and establishment of the order in which events and outcomes occur. However, survivorship bias in longitudinal mental health surveys suggest that longitudinal samples may be non-representative of population-level mental health. While unable to determine causation, cross-sectional studies can more rapidly generate data, and our data provide further evidence that cross-sectional data may be more reliable for the assessment of population-level prevalences of adverse mental health symptoms at a given timepoint.^35^ For future study designs, researchers could consider implementing a planned missing data design^36^ to benefit from the strengths of these study designs while minimizing associated biases.

Strengths of this analysis include four timepoints to assess response bias, high initial response (61.7%) and retention (39.6% of respondents completed three of four surveys) rates, utilization of clinically validated screening instruments, and implementation of quota sampling and survey weighting to improve sample representativeness by national estimates for gender, age, and race/ethnicity. Moreover, multiple types of survivorship bias were assessed, including differential demographic attrition and demographic-adjusted assessment of both initial mental health as well as odds of participation in follow-up surveys based on changes to mental health over the initial two surveys. Finally, bias was assessed both cross-sectionally and longitudinally. The findings in this report are also subject to limitations. First, while this analysis focused on survivorship bias, these data may be subject to other biases, including recall and response biases^37,38^; however, quota sampling and survey weighting were employed to reduce demographic-related response bias. Second, though strategies were used to improve sample representativeness, and this Internet-based survey sample should represent the adult U.S. population by gender, age, and race/ethnicity, it may not fully represent all U.S. adults, especially with regards to Internet access. Third, April-2020 respondents who did not respond to invitations to complete surveys in either May-2020 or June-2020 were not invited to complete September-2020 surveys, so these respondents did not have the opportunity to complete September-2020 surveys. However, after having declined two successive invitations, it is unlikely that a substantial number of these respondents would have completed September-2020 surveys. Finally, a portion of the sample oversampled from the New York City and Los Angeles metropolitan areas. However, all 50 states and Washington D.C. were represented, and this analysis was not designed to produce national population estimates for adverse mental health symptoms.

Longitudinal survey-based assessment of mental health is a useful and widely used research method that can provide important insights gained from monitoring the same participants over time. However, our data demonstrate that analysing mental health trends among only individuals who consistently respond to longitudinal mental health surveys can lead to overly optimistic interpretations of mental health trends by excluding individuals who less frequently respond to follow-up survey invitations. Survivorship bias assessment should therefore be among bias assessments^38-41^ applied before conclusions based on repeated assessments from participants in a longitudinal study are generalized. These data have critical implications for the design of future studies and interpretation of data from published papers and ongoing studies with longitudinal study designs, both during and beyond the COVID-19 pandemic.

## Data Availability

All relevant data supporting the findings in this study are available from the corresponding author upon reasonable request.

## Additional sections

### Ethics approval

The protocol was approved by the Monash University Human Research Ethics Committee (MUHREC) (ref. no. 24036). This activity was also reviewed by the U.S. Centers for Disease Control and Prevention (C.D.C.) and was conducted consistent with applicable federal law and C.D.C. policy: 45 C.F.R. part 46, 21 C.F.R. part 56; 42 U.S.C. Sect. 241(d); 5 U.S.C. Sect. 552a; 44 U.S.C. Sect. 3501 et seq.

## Acknowledgements

We thank all survey respondents, along with Laura K. Barger, Ph.D. (Brigham & Women’s Hospital, Harvard Medical School), Rebecca Robbins, Ph.D. (Brigham & Women’s Hospital, Harvard Medical School), Elise R. Facer-Childs, Ph.D. (Monash University), and Matthew D. Weaver, Ph.D. (Brigham & Women’s Hospital, Harvard Medical School) for their contributions to the initial survey instrument for The COPE Initiative.

## Author contributions

M.É.C., C.A.C., S.M.W.R., and M.E.H. designed the study. M.É.C. and J.F.W. conceived the manuscript. M.É.C. worked with Qualtrics, LLC research services to administer the survey, and analysed the data with guidance from J.F.W. M.É.C. created the table and all figures. M.É.C. wrote the first paper draft. All authors provided critical input and revisions to the paper. S.M.W.R. and M.E.H. supervised.

## Disclaimer

This manuscript reflects the views of the authors and does not necessarily represent the official position of the Centers for Disease Control and Prevention.

## Ethical Standards

The authors assert that all procedures contributing to this work comply with the ethical standards of the relevant national and institutional committees on human experimentation and with the Helsinki Declaration of 1975, as revised in 2008.

## Availability of Data and Materials

All relevant data supporting the findings in this study are available from the corresponding author upon reasonable request.s

